# Therapeutic Restoration of Systemic Multiomic Responses by Transglutaminase 2 inhibitor in Celiac Disease: A Gluten Challenge Approach

**DOI:** 10.1101/2025.10.31.25339231

**Authors:** Valeriia Dotsenko, Hana Hien Le, Sonja Rajić, Robert Moulder, Jalmari Kettunen, M. Karoliina Hirvonen, Alex M Dickens, Tuulia Hyötyläinen, Bernhard Tewes, Timo Zimmermann, Ralf Mohrbacher, Tomi Suomi, Terho Lehtimäki, Riitta Lahesmaa, Matej Orešič, Laura L. Elo, Emma Raitoharju, Detlef Schuppan, Markku Mäki, Keijo Viiri, CEC-3 Investigators

## Abstract

**Background and Aims:** Celiac disease (CeD) is an autoimmune disease triggered by dietary gluten in genetically predisposed individuals. Deamidation of gluten peptides by the CeD autoantigen and enzyme transglutaminase 2 (TG2) is central to the pathogenesis of CeD. Inhibition of TG2 with the specific inhibitor ZED1227 effectively prevents gluten-induced histological damage in CeD patients. Here we aimed to explore the systemic plasma lipidomic, proteomic and DNA methylomic changes in ZED1227-treated CeD patients undergoing a gluten challenge.

**Methods:** Individuals with CeD on a long-term gluten-free diet (GFD) underwent a 6-week gluten challenge combined with daily 100mg ZED1227 drug (PGCd, n = 28) or placebo (PGCp, n = 19). Samples were collected at baseline (GFD) and post-gluten challenge (PGC). Mass spectrometry-based lipidomic and proteomics profiling, along with genome-wide DNA methylation analysis, were applied to plasma samples matched with duodenal histology. Comparative analyses were performed between the groups, with adjustment for BMI, age, sex, and country of origin.

**Results:** Significantly different gluten-induced plasma lipidomic changes were detected between GFD vs. PGCp and between GFD vs. PGCd, with 46 lipids differentially expressed in the placebo group and 6 in the drug group suggesting that the ZED1227 normalized gluten-induced lipidomic changes in plasma. Changes in medium-chain fatty acylcarnitines (CARs), particularly CAR 10:1 and CAR 9:0 correlated with kidney function which decreased significantly in PGCp. Glomerular filtration rate and plasma creatinine were restored with ZED1227. Drug treatment revealed consistent patterns suggesting normalization of the proteome and DNA methylome indicating that ZED1227 attenuated the systemic responses to gluten challenge.

**Conclusions:** These findings provide evidence that ZED1227 can significantly prevent the gluten-induced CeD-associated systemic changes in the plasma.

## INTRODUCTION

Celiac disease (CeD) is a small intestinal autoimmune enteropathy triggered by dietary gluten in genetically susceptible individuals. CeD is a complex, multifactorial disease marked by a T-cell mediated immune response to gluten peptides which are resistant to complete digestive degradation due to their high proline and glutamine content.^1^ Genetic susceptibility plays a key role in CeD pathogenesis, with the human leukocyte antigen (HLA) class II molecules-particular HLA-DQ2 and HLA-DQ8 heterodimers-being the primary genetic determinants. Globally, CeD affects about 1.4% of the population based on serology with biopsy-confirmed cases at 0.7%, with rising prevalence.^2^ This highlights the critical need for early detection and effective dietary management of CeD.^3^

Patients, both children and adults, can exhibit a spectrum of inflammatory enteropathy, clinically being asymptomatic or presenting as a severe malabsorptive syndrome, often with extraintestinal manifestations with or without gastrointestinal symptoms.^4,5^ Certain molecular mechanisms of CeD remain elusive despite progress in genetic and immunological research.^6,7^ In patients without or delayed therapy, i.e., the gluten-free diet (GFD), CeD can lead to high morbidity and even mortality, contributing to complications such as neurological disorders,^8–10^ osteoporosis,^11,12^ kidney diseases,^13–15^ autoimmune endocrine diseases,^16^ especially type 1 diabetes and autoimmune thyroiditis,^17^ and various malignant complications.^18–20^ Even with adherence to the GFD, up to 30% of patients continue to suffer from CeD-related symptoms^21^, often confirmed by histological signs of inflammation and villous atrophy.^22^

Although strict adherence to the GFD usually improves major intestinal and some extraintestinal manifestations of CeD, maintaining this diet is difficult due to inadvertent ingestion of traces of gluten that are present in most prepared foods, resulting in continuing symptoms,^23^ long-term small intestinal mucosal injury,^24^ adversely affecting health and quality of life.^25,26^ In light of these challenges, the past decade has seen significant research efforts directed toward developing pharmacological treatments for CeD.^27,28^ A promising advance in this field is ZED1227, an oral, first-in-class selective inhibitor of transglutaminase 2 (TG2), which has shown safety and efficacy in Phase 1 clinical trials.^29^ A recent Phase 2 proof-of-concept trial revealed that ZED1227 could largely prevent gluten-induced intestinal mucosa injury while also decreasing symptoms and improving the quality of life for CeD patients.^30,31^

There has been a growing interest in understanding metabolic and lipidomic alterations in CeD. Recent efforts have focused on identifying metabolic alterations in the serum of potential adult^32^ and pediatric^33,34^ CeD patients, including signatures that may precede the clinical manifestations. The GFD has been shown to improve metabolic^35^ and lipid profiles, which are particularly affected by CeD.^36^ However, there is a lack of research on the lipidomic markers in the blood of CeD patients undergoing a gluten challenge, especially when combined with effective drug treatment.

Recent studies of CeD biopsies suggest that proteomic data may aid in diagnosing morphological changes in the duodenal mucosa^37,38^. Furthermore, a plasma proteomic study has suggested differentially expressed proteins that may serve as potential diagnostic biomarkers for CeD.^39^ Importantly, only one study has suggested that blood-based DNA methylation biomarkers may be useful in detecting chronic gastrointestinal disorders such as CeD.^40^ While gluten exposure triggers an immediate systemic cytokine response, most notably an increase in interleukin-2 (IL-2), ^41,42^ there are currently no published studies investigating the long-term effects of gluten-induced changes in the plasma proteome and DNA methylome in conjunction with pharmacological treatment in CeD.

Using our well-defined cohort of CeD patients in remission challenged with gluten over six weeks and treated with ZED1227 vs placebo, we sought to investigate the systemic plasma lipidomic, proteomic and DNA methylomic signatures to identify potential early systemic markers of gluten exposure, evaluate the drug’s efficacy in reducing the systemic response to gluten, and explore the broader implications of this drug treatment.

## RESULTS

### Plasma lipidomic profiling reveals broad structural shifts following gluten challenge

To characterize lipidomic alterations induced by gluten exposure, plasma lipid profiles were measured in CeD patients receiving placebo or ZED1227 after 6 weeks of gluten challenge (PGC) (Fig. S1). A total of 308 lipid species, spanning six structural categories, were identified and quantified. Log-transformed intensity distributions were consistent across all sample groups, indicating high data quality (Fig. 1A, B).

**Figure 1.**
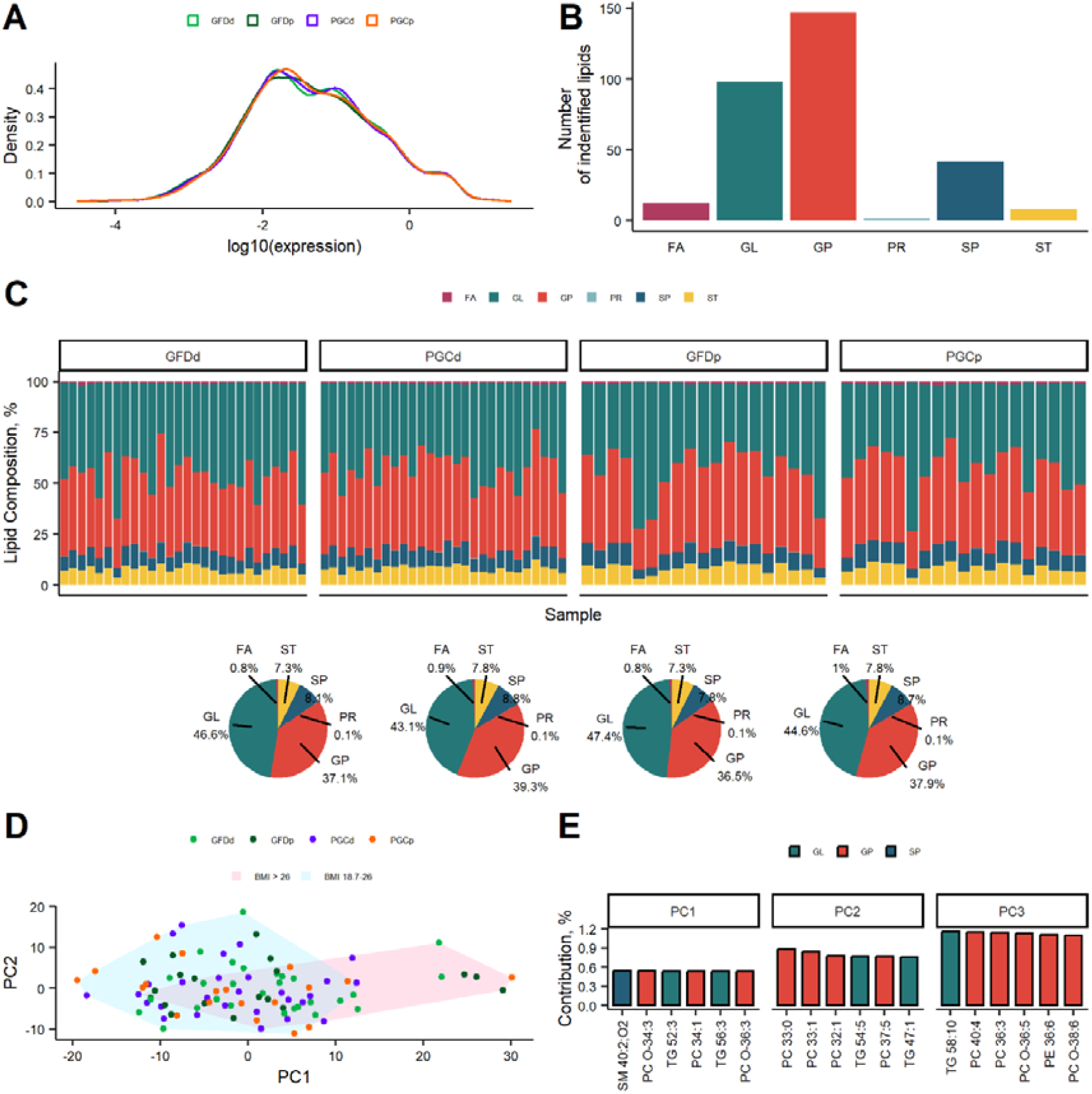
Lipidomic Analysis and Principal Component Distribution in Celiac Patient Plasma Samples. **A)** The density distribution of detected lipid expression after log10 transformation reveals a uniform distribution across all four sample groups: total samples (n = 94), GFDd (n = 28), GFDp (n = 19), PGCd (n = 28), and PGCp (n = 19). **B)** Barplot illustrates the number of identified lipids (n = 308) in the plasma of celiac patients, categorized by their structural type: Fatty Acyls (FA), Glycerolipids (GL), Glycerophospholipids (GP), Prenol Lipids (PR), Sphingolipids (SP) and Sterol Lipids (ST). **C)** The composition of lipids by structural category is presented in barplots for individual patients (top) and collectively for patient groups (bottom circular diagram). GP, GL and SP were identified as the primary lipid components in plasma. Notably, there was a decrease in GL and an increase in GP in both the placebo and drug PGC groups. **D)** Principal Component Analysis (PCA) scores plot demonstrates that the first and second principal components (PC1 and PC2) account for 32.7% and 11.1% of the variance, respectively. Sample groups, including GFDd, GFDp, PGCd, and PGCp, are represented by green, dark green, blue, and orange circles, respectively. Shaded areas in pink and blue represent samples with BMI values greater than 26 and between 18.7 and 26, respectively. Although there’s no clear separation of the samples by groups, there’s a notable correlation between PC1 and BMI. **E)** This panel highlights the top contributors to PC1, PC2, and PC3. A majority of these contributors belong to the GP and GL categories.

Lipid composition was further examined at the structure category level. While individual-level variability was present, no extreme outliers were detected (Fig. 1C, top level). The major lipid components in plasma were identified as Glycerophospholipids (GP), Glycerolipids (GL), and Sphingolipids (SP) (Fig. 1C). Following gluten challenge, the proportion of GP increased in both ZED1227-treated (PGCd, from 37.1% to 39.3%) and placebo-treated (PGCp, from 36.5% to 37.9%) groups, while GL proportion decreased in both PGCd and PGCp from 46.6% and 47.4% to 43.1% and 44.6%, respectively (Fig. 1C, bottom panel), suggesting a consistent gluten-induced shift in lipid composition.

On principal-component analysis (PCA) there was no difference between groups based on diet or treatment. However, stratification by body mass index (BMI) revealed a clear separation, with individuals having a BMI > 26 clustering distinctly along PC1 (Fig. 1D). PC1 accounted from 32.7% of the total variance and was significantly associated with BMI (p < 0.001). The top contributors to PC1 are shown in Figure 1E, with the majority of lipids belonging to the GP and GL class. When compared to BMI, the trend was consistent in both the GFDp and PGCp groups (Fig. S2), indicating that the BMI-lipidome relationship was maintained regardless of gluten exposure. Additionally, PC3 was significantly correlated with sex and country of residency during the drug treatment, while no significant correlations were observed between the first three principal components and age (Table 2).

### Gluten challenge induced differential lipid expression is attenuated by ZED1227 treatment

To identify lipids significantly altered by gluten exposure and drug treatment, we performed differential expression analysis with patient characteristics such as age and sex. Considering the influence of patient characteristics on PC analyses (Fig.1D, Table 2), these variables were included as covariates in the fitted model to determine differential lipid expression. The greatest number of differentially expressed lipids (DELs, n = 46) was detected in the PGCp vs. GFDp comparison (Fig. 2A), the two most abundant structural categories being GP (21 DELs) and GL (18 DELs) (Fig. 2E and Supplemental Data 1). Among the 46 DELs, the findings showed upregulation of two medium-chain acylcarnitines-CAR 10:1 (9-decenoylcarnitine) and CAR 9:0 (Nonanoylcarnitine) (Fig. 2C, middle panel).

**Figure 2.**
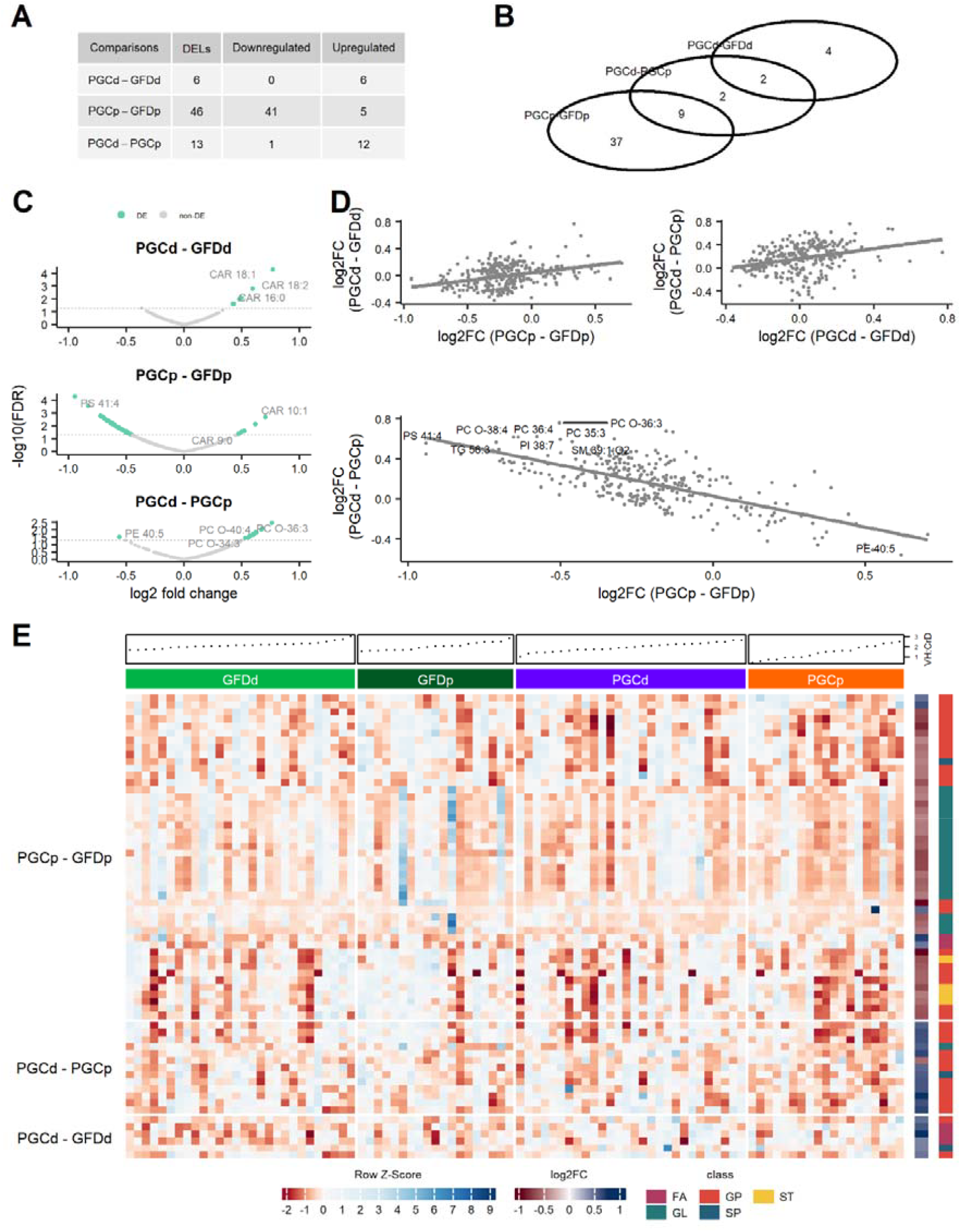
Changes of plasma lipidomic profiles of celiac patients after gluten challenge combined with placebo or ZED1227 treatment. **A)** Table showing the number of differentially expressed lipids (DELs) in the indicated comparisons: 6 for PGCd vs GFDd, 46 for PGCp vs GFDp, 13 for PGCp –vs PGCd; total samples (n = 94), GFDd (n = 28), GFDp (n = 19), PGCd (n = 28), and PGCp (n = 19). **B)** Venn diagram illustrating the number of DELs that are shared in the comparison of PGCp vs PGCd and PGCp vs GFDp (9 DELs); PGCp vs PGCd and PGCd vs GFDd (2 DELs). **C)** Volcano plot representations comparing DELs. Green dots indicate statistically significant DELs with a p-value ≤ 0.05. The dashed horizontal line represents the p-value threshold. CAR - fatty acyl carnitines; CAR 18:2 – octadecadienylcarnitine; CAR 18:1 – octadecenoylcarnitine; CAR 16:0 – palmitoylcarnitine; CAR 10:1 – 9-decenoylcarnitine; CAR 9:0 – nonanoylcarnitine; PS – diacylglycerophosphoserines; PE – diacylglycerophosphoethanolamines; PE 40:5 - PE(18:1e/22:4), PC O--glycerophosphocholines. **D)** Correlation profiles of all identified lipids (*n* = 308) log2FC between PGCp vs GFDp and PGCd vs GFDd, PGCd vs GFDd and PGCd vs PGCp show consistent directionality of lipid expression changes. In contrast, the PGCp vs GFDp and PGCd vs PGCp correlations show opposite directionality of lipid expression. **E)** Heatmap depicting the expression of all detected DELs in tested comparisons across all samples. Lipids in rows are grouped by group comparisons, and samples are in ranking order of increase in histological villus height vs crypt depth (VH:CrD), as depicted in the charts above the heatmaps. DELs are categorized by their structural type: Fatty Acyls (FA), Glycerophospholipids (GP), Sterol Lipids (ST), Glycerolipids (GL), and Sphingolipids (SP).

When comparing the PGCd vs. GFDd, only 6 DELs were found, with no overlap with the DELs in the PGCp vs. GFDp comparison (Fig. 2B), suggesting that the drug largely normalized gluten-induced lipidomic changes in plasma. In the PGCd vs. GFDd comparison, lipids belonging to 3 structural categories were detected – FA (3 DELs), GP (2 DELs), and SP (1 DEL) (Fig. 2E and Supplemental Data 1). Notably, half (or 25% of all identified carnitines) of the DELs are long-chain carnitines: CAR 18:2, CAR 18:1, and CAR 16:0 (Fig. 2C, upper panel).

In the PGCd vs. PGCp comparison, lipids from three structural categories were detected – GP (11 DELs,), GL (1 DEL), and SP (1 DEL), including nine that overlapped with the PGCp vs. GFDp comparison (Fig. 2B, 2C, and Supplemental Data 1). Crucially, these nine DEL lipids exhibit opposite regulation patterns between the two comparisons (Fig. 2D). Interestingly, when plotting all detected lipids (Fig. 2D, lower panel) there was a clear trend: lipids that increased or decreased after gluten challenge within individuals tended to show opposite direction in the interindividual comparison between PGCd vs. PGCp, suggesting that ZED1227 treatment normalizes gluten-induced lipid changes.

### ZED1227 inhibits gluten-induced alterations in systemic plasma proteomic profiles and DNA methylation landscapes

The discovery that ZED1227 can restore the gluten-induced plasma lipidomic changes prompted us to take an extended multiomics approach to investigate this further. Differential protein expression was tested between baseline and gluten exposure among placebo-treated patients, PGCp vs GFDp, as well as post-exposure between drug- and placebo-treated, PGCp vs PGCd, patients. After multiple testing corrections, no protein reached statistical significance in either of the two comparisons (adjusted p > 0.05, data not shown). However, when the expression changes of all proteins from both comparisons were assessed, a positive correlation of 0.55 was observed (p < 10^-15^, Fig. 3A). This suggests that treatment with ZED1227 alters the patient’s plasma proteome in the same direction as a gluten-free diet, when compared to gluten-challenged state under placebo treatment.

**Figure 3.**
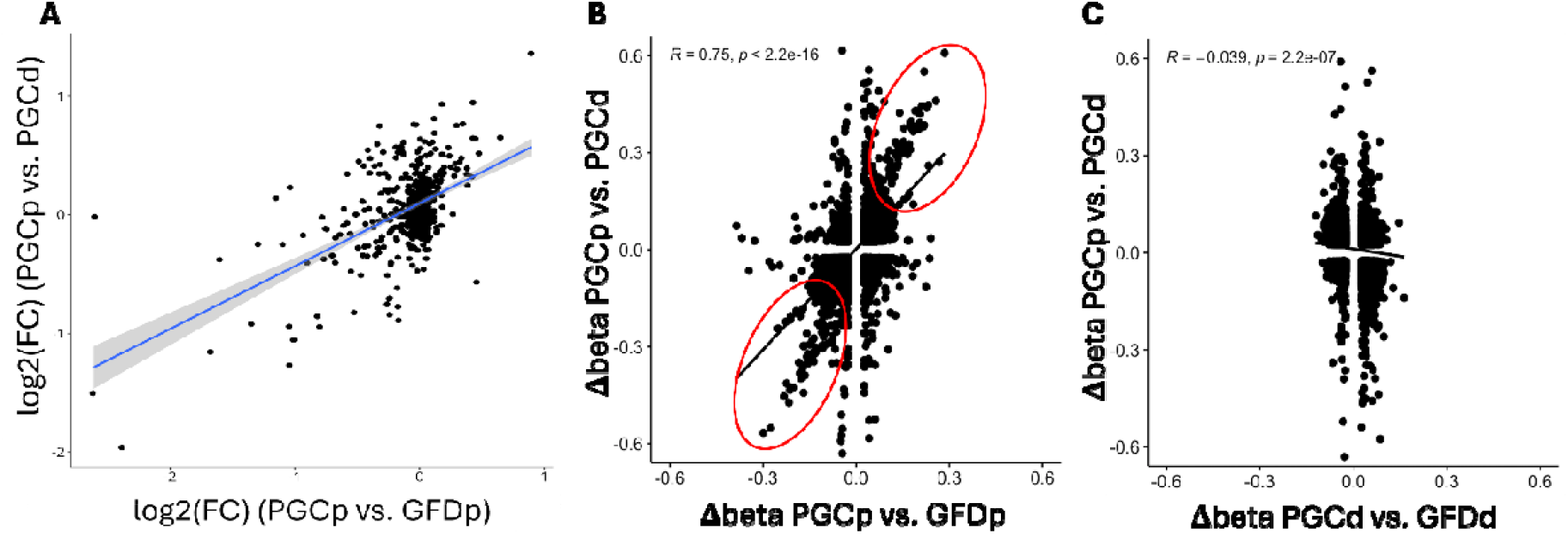
Systemic proteomic and epigenetic restoration with ZED1227. **A)** Correlation between log_2_ fold changes of comparisons PGCp vs GFDp and PGCp vs PGCd in the proteomics data (Pearson correlation coefficient 0.55, p < 10^-15^). Each data point represents a protein (n = 512). Shading indicates the 95 % confidence interval of the fitted regression line. **B)** CpG sites having changes in methylation more than 0.025 beta units (Δβ) are shown. Red circles indicate the strong correlation in comparison between intraindividual Δβ’s in PGCp vs. GFDp and interindividual Δβ’s in PGCp vs. PGCd. **C)** No such correlation was detected when intraindividual Δβ’s in PGCd vs. GFDd was compared to interindividual Δβ’s in PGCp vs. PGCd.

Given the observed changes in both lipidomic and proteomic analysis, we investigated whether these were accompanied by epigenetic modifications via DNA methylation. DNA methylation analysis of approximately 800,000 CpG sites from peripheral blood revealed no statistically significant differences (FDR < 0.05) between baseline and gluten exposure in the full cohort, or within placebo- or drug-treated subgroups. To explore broader methylation trends beyond stringent significance thresholds, CpG sites exhibiting an absolute change in methylation (Δβ) greater than 0.025 were examined, independent of p-value. This approach identified a subset of CpG sites displaying concordant methylation shifts in both the placebo group (GFD vs. PGC) and between PGC groups (placebo vs. ZED1227). The magnitude of methylation change at these CpG sites was strongly correlated between the two comparisons (Pearson’s R = 0.75), suggesting that gluten-induced epigenetic alteration in the placebo group is partially reversed or attenuated in the presence of ZED1227 (Fig. 3B). In contrast, no such relationship was observed when comparing the placebo vs. drug treated groups at PGC with the GFD vs. the drug-treated PGC group, which showed only a weak negative correlation with R = −0.039 (Fig. 3C). These findings suggest that ZED1227 may dampen systemic epigenetic responses to gluten challenge, particularly at CpG sites altered in untreated individuals.

### Carnitine-associated lipid profiles correlate with kidney function during gluten challenge and TG2 inhibitor treatment

Enrichment analysis of lipid categories revealed fatty acyls (FA) consisting of fatty acyl carnitines (CARs) as a key group affected by both gluten and drug treatment. FAs were upregulated in PGCp vs. GFDp and PGCd vs. GFDd, and downregulated in PGCd vs. PGCp, highlighting their role as markers of metabolic response (Fig. 4A). Notably, long-chain CARs, including CAR 18:2, CAR 18:1, and CAR 16:0 were upregulated in the PGCd in comparison with its baseline (GFDd) while medium-chain CARs (CAR 10:2 and CAR 9:0) increased in PGCp (Fig. 2C). However, the total concentration of CARs detected did not appear to differ among the patient groups (Fig. 4B). When CARs were categorized by the length of their fatty acid chains the expression levels of all lengths were comparable, except for L-carnitine (0) and acetylcarnitine (2) (Fig. 4C).

**Figure 4.**
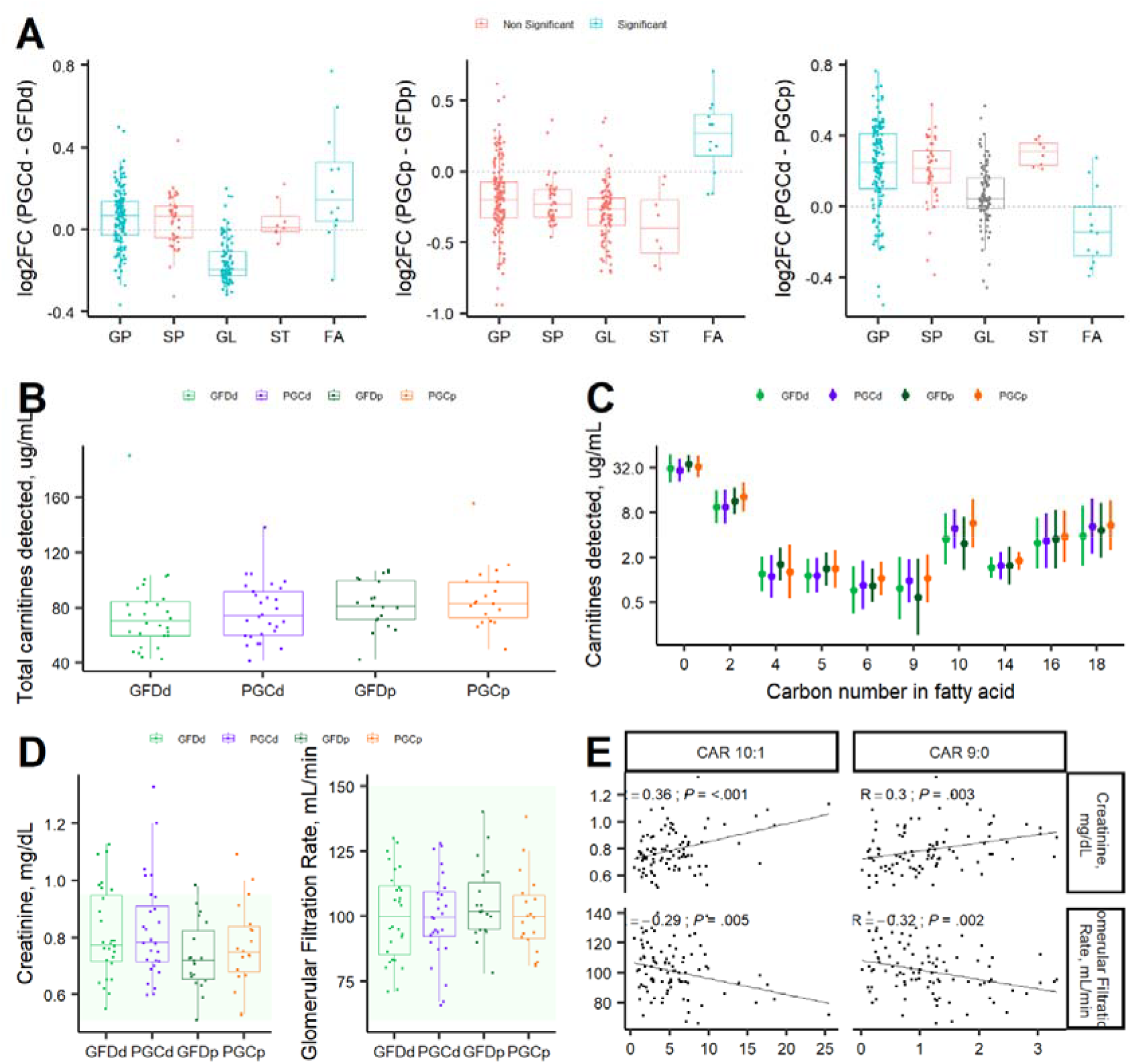
CAR 10:1 and CAR 9:0 correlate with indicators of kidney functions in celiac patients before and after the gluten challenge. **A)** Enrichment analysis. Lipids were ranked by their fold change and then grouped by their structural categories. Fatty acyls (FA, consisting of fatty acyl carnitines) and glycerophospholipids (GP) were over-represented in both PGCp and PGCd groups. Fatty acyls (FA), glycerophospholipids (GP), sterol lipids (ST), sphingolipids (SP), glycerolipids (GL), and prenol lipids (PR). **B**) Expression of total carnitines detected in the plasma of celiac patients, divided by groups: total samples (n = 94), GFDd (n = 28), GFDp (n = 19), PGCd (n = 28), and PGCp (n = 19). **C)** Carnitines are expressed by the length of their fatty acid. Fatty acylcarnitines with chain lengths 10 and 9 were differentially expressed in the PGCp-GFDp comparison. **D)** Levels of Creatinine in mg/dL in plasma and the calculated Glomerular filtration rate in mL/min for celiac patients. The green shading indicates the normal range for these laboratory tests (0.51 - 0.95 mg/dL for plasma Creatinine and 60 – 150 mL/min for glomerular filtration Rate). The figure includes the results of a two-sided paired t-test for p-value calculations, highlighting statistically significant findings (*P* < .05). **E)** The correlation of plasma creatinine and glomerular filtration rate indicators with CAR 10:1 and CAR 9:0 is statistically significant. Pearson correlation coefficient is shown, and P-values less than 0.05 are considered to be significant. CAR 10:1 – 9-decenoylcarnitine; CAR 9:0 – Nonanoylcarnitine.

While the differential expression of CARs provides insight into metabolic responses to treatment, carnitine blood levels have also been shown to have variation in different diseases, such as liver cirrhosis of various etiologies,^43,44^ renal disease,^45^ Crohn’s disease,^46^ and CeD.^47^ In order to understand the clinical implications of these findings, we checked routine laboratory test results available for our study cohort.

To assess the clinical significance of CAR alteration, we analysed kidney function, as indicated by plasma creatinine levels and glomerular filtration rate (GFR). Patients in the placebo group exhibited a tendency towards decreased kidney function, as indicated by increase in plasma creatinine (from 0.73 ± 0.12 to 0.77 ± 0.15 mg/dL, P = .01) and a decrease in GFR (from 105.1 ± 14.8 to 100.8 ± 15.1 mL/min, P = 0.006) following the 6-week gluten challenge, although values remained within normal ranges (Fig. 4D). In contrast, no significant kidney function changes were observed in the drug-treated group (PGCd) with no significant reduction in kidney function (GFR mean ± SD in mL/min: at GFDd 100.3 ± 17.1, at PGCd 99.6 ± 16.5, P = 0.72; creatinine mean ± SD in mg/dL: at GFDd 0.81 ± 0.16, at PGCd 0.83 ± 0.18, P = 0.36) (Fig. 4D).

Correlation analysis demonstrated that CAR 10:1 and CAR 9:0 negatively correlated with GFR (R = −0.29, P = 0.005 and R = −0.32, P = 0.002, respectively) and positively with plasma creatinine levels (R = 0.36, P < 0.001 and R = 0.30, P = 0.003, respectively) (Fig. 4E), reinforcing their potential role as metabolic indicators of gluten-induced renal stress. CARs identified as DELs in the comparison between PGCd and GFDd groups showed no significant correlation with plasma creatinine levels, with the exceptions being CAR 16:0 and CAR 18:1, which were found to correlate with GFR (Fig. S3).

## DISCUSSION

In celiac, gluten exposure initiates an immune response that disrupts gut homeostasis and leads to intestinal damage. This immune activation can cause systemic changes, including alterations in lipid metabolism detectable in the blood. Lipidomic analyses have shown that patients with active CeD exhibit altered serum or plasma lipid profiles, with changes in levels of glycolipids (GLs), in particular triglycerides (TGs)^34^ and diacylglycerols (DGs),^48^ phosphatidylcholines (PCs),^33,34^ cholesterol derivatives,^34^ and fatty acyls (FA).^48^ Such alterations are already evident in infants even before gluten exposure, for example, changes in phospholipids levels^33^ and upregulation of triacylglycerols.^34^ Some studies detected no differences in systematic levels of fatty acids in newly diagnosed CeD.^49^ However, there are no published results on the lipidomic changes in adult CeD patients post-gluten challenge, or with/without effective drug therapy.

In the present study, we have observed distinct plasma lipidome profiles in CeD patients who underwent a gluten challenge, with and without the TG2 inhibitor ZED1227. Notable differences emerged in both cohorts receiving gluten, reflecting the specific treatments administered, placebo or TG2 inhibitor drug ZED1227. Out of a total of 1004 lipids detected, 308 were successfully identified. Among these identified lipids, 46, accounting for approximately 15%, demonstrated differential expression in the placebo group following a gluten challenge. Specifically, 41 lipids decreased, while 5 increased. Notably, the fatty acyl (FA) category was significantly enriched in comparison between the gluten challenged and placebo-treated group (PGCp) and the baseline gluten-free diet placebo group (GFDp). FAs, particularly eicosanoids, were previously found to be upregulated in the plasma of children with CeD compared to their non-celiac siblings.^48^ However, another study did not observe changes in plasma fatty acids in children with newly diagnosed CeD.^49^ These differing results might be attributed to methodological variances and differences in the studied cohorts, making direct comparisons with our findings difficult.

Interestingly, gluten challenged patients treated with ZED1227 showed significantly less differentially expressed lipids, as only six lipids were differentially expressed in the GFDd vs. PGCd comparison (Fig. 2A). Importantly, none of the lipid alterations evoked in gluten challenge appeared in patients treated with ZED1227 (Fig. 2B). This suggests that attenuation of gluten-induced mucosal damage of ZED1227 can be also detected in lipidomic changes in plasma. Additional proof for this comes as correlation profiles of all detected lipids (n = 308) when PGCp vs. PGCd and PGCp vs. GFDp comparisons show opposite directionality of lipid expression, which strongly suggests that ZED1227 can inhibit gluten-induced lipidomic changes (Fig. 2D).

Besides lipidomics, proteomics analysis, although showing no individual proteins with statistically significant differential expression after multiple testing correction, revealed global expression patterns suggesting the ZED1227 modulates the plasma proteome in a manner similar to a gluten-free diet (Fig. 3A). This supports that TG2 inhibition influences systemic immune and metabolic responses. Additionally, epigenetic profiling did not identify significant site-specific DNA methylation changes after gluten challenge or drug treatment. However, analysis of CpG sites with moderate methylation shifts indicated that gluten-induced epigenetic alterations in untreated patients were partially reversed or attenuated by ZED1227 (Fig. 3B&C). These findings highlight that TG2 inhibition reduced systemic epigenetic responses to gluten exposure, potentially contributing to its therapeutic efficacy in CeD patients.

An exploratory finding in this study was a reduction in kidney function tests following a small amount (3g) and only a 6-week-long gluten challenge. The values of the gluten-induced changes in plasma creatinine and GFR remained within normal limits but were still clearly corrected upon ZED1227 treatment, i.e., when blocking the downstream pathogenetic effects induced by gluten in CeD patients. In fact, after years and decades of gluten ingestion, untreated CeD has been linked in clinical and epidemiological studies to an increased risk of kidney diseases.^14,50,51^ Our preliminary and incidental result suggest a direct gluten effect on worsening kidney function, supporting the link between kidney diseases and gluten-induced extraintestinal manifestations in CeD. Further, the kidney function tests (in the PGCp group) correlated with plasma levels of medium-chain acylcarnitines, CAR 9:0, and CAR 10:1. In fact, increased plasma concentrations of pelargonylcarnitine (CAR 9:0)^52^ and decanoylcarnitine (CAR 10:0)^53^ have earlier been reported in patients with impaired kidney function. In the kidney, carnitine and its precursors are efficiently reabsorbed to minimize urinary loss.^54^ It has been proposed that acylcarnitines may serve as metabolomic markers for chronic kidney disease.^55^ Several studies have found an inverse association between long-chain acylcarnitines,^53^ as well as short-chain^56,57^ and medium-chain acylcarnitines,^57^ and reduced estimated glomerular filtration rate (GFR). Further studies are warranted to show whether these acylcarnitines could serve as early biomarkers for the systemic effects of gluten ingestion in CeD patients.

Carnitine plays a crucial role in energy metabolism by facilitating the transport of long-chain fatty acids into mitochondria for β-oxidation. Blood plasma levels of carnitine are maintained through dietary intake, endogenous synthesis, renal reabsorption, and cellular uptake.^58^ CeD can lead to secondary carnitine deficiency.^59^ Significantly lower serum total carnitine concentrations have been observed in patients compared to those on a gluten-free diet (GFD) and non-CeD controls.^60^ Additionally, various acylcarnitines (CAR) have been reported to decrease in CeD patients on a GFD compared to healthy individuals,^61^ a condition often linked to intestinal inflammation and resulting malabsorption syndromes. Our study, however, did not detect deficiencies in plasma concentrations of L-carnitine and short-chain CARs (C1-C4). This could be attributed to our cohort of well-controlled celiac patients at baseline, and the relatively short duration of the gluten challenge they underwent. Furthermore, while an increase in Isobutyryl-L-carnitine (CAR 4:0) levels has been observed in the plasma of pediatric CeD patients showing disease progression,^62^ this trend was not evident in our study’s patients.

We acknowledge certain limitations of the current study. The sample size is relatively low, comprising only 47 CeD patients; however, the patients have been broadly characterized clinically, histologically and regarding laboratory parameters, which we consider adequate for analysis using robust statistical methods. Even if the gluten challenge duration was relatively short—six weeks—with a daily intake of an intermediate amount of gluten (3g vs an average daily intake of 10-15g in the non-celiac population), we observed significant lipidomic alterations upon gluten challenge, which were significantly attenuated with TG2 inhibitor treatment. Similar trends were also observed when monitoring proteomic and DNA methylomic changes (Fig. 3). The identified differentially expressed lipid species can provide a foundation for future research aimed at establishing novel biomarkers reflecting dietary effects of gluten. To conclude, our integrative multi-omics approach provides a holistic view of the molecular landscape in CeD patients undergoing gluten challenge and supports the notion that TG2 inhibitor treatment has also beneficial systemic effects.

## Supporting information

Lict of CEC-3 investigators

## ACKNOWLEDGEMENTS

We thank the patients who participated for making this study possible. We also thank the expert staff for their participation in the sample collection. This work was supported by the Research Council of Finland (Grant no. 370828), the Finnish Cultural Foundation, Dr. Falk. Pharma GmbH, Mary och Georg C. Ehrnrooths Stiftelse, and the State funding for university-level health research, Tampere University Hospital, Wellbeing services county of Pirkanmaa / Project No. T63474 and T66984. SR acknowledges support from Yrjö Jahnsson foundation and Päivikki and Sakari Sohlberg foundation. TL acknowledges Research Council of Finland (Grant no. 356405), Finnish Foundation for Cardiovascular Research and Juho Vainio Foundation. RL acknowledges support from the Research Council of Finland (Grant no. 331793 and 329277), RL, LLE and MO acknowledge the support for InFLAMES Flagship Programme from Research Council of Finland (Grant no. 337530). LLE acknowledges support from the Research Council of Finland (Grant no. 341342, 364700). ER acknowledges support from the Research Council of Finland (Grant no. 338395). DS acknowledges CeD-related research support from Collaborative Research Centre grant TRR 355/1 (490846870) project B08, and EU-BMBF project ImmunoSafe-CeD (01EA2205B). Research is also supported by Biocenter Finland. The funding sources played no role in the design or execution of this study or in the analysis and interpretation of the data.

## ETHICS APPROVAL

TUKIJA dnro 223/06.00.01/2017, EudraCT 2017-002241-30 for Dr. Falk Pharma funded clinical trial. The study was conducted with de-identified data of the participants who had consented to the use of their anonymized data in research.

## DATA AVAILABILITY

The mass spectrometry discovery proteomics data have been deposited to the ProteomeXchange Consortium via the PRIDE^63^ partner repository with the dataset identifier PXD069597.

## AUTHOR CONTRIBUTIONS

KV, MM and VD conceptualized the study. KV, VD and HHL drafted the manuscript. VD, HHL, SR, RM, JK, MKH, AMD, ER and KV. performed investigation, data analysis and figure generation. TH, TS, TL, RL, MO, LLE and KV, provided resources and data curation. BT, TZ, RM, and DS assisted in the logistics of data collection and results interpretation. KV and MM acquired the funding. All authors read and approved the final paper.

## COMPETING INTEREST

Valeriia Dotsenko, none to declare.

Hana Hien Le, none to declare.

Sonja Rajic, none to declare.

Robert Moulder, none to declare.

Jalmari Kettunen, none to declare.

M. Karoliina Hirvonen, none to declare.

Alex M Dickens, none to declare.

Tuulia Hyötylänen, none to declare.

Bernhard Tewes, employee of Dr. Falk Pharma GmbH.

Timo Zimmermann, employee of Dr. Falk Pharma GmbH.

Ralf Mohrbacher, employee of Dr. Falk Pharma GmbH.

Tomi Suomi, none to declare.

Terho Lehtimäki, none to declare.

Riitta Lahesmaa, none to declare.

Matej Oresic, none to declare.

Laura Elo, none to declare.

Emma Raitoharju, none to declare.

Detlef Schuppan, consultant for Falk Pharma, Chugai, Immunic, Sanofi and Tillots.

Markku Mäki, has received during the last three years Management/Advisory Affiliation fees from Dr. Falk Pharma, Interlude Biopharma, Entero Therapeutics, Calypso Biotech, ImmunogenX, and Immunic; holds patent licensed to Labsystems Diagnostics from where MHT has received royalties via Tampere University Hospital.

Keijo Viiri, none to declare.

## MATERIALS AND METHODS

### Patient recruitment, study design, sample collection and processing

This study utilized plasma-EDTA samples from a multi-site, double-blind, randomized, placebo-controlled trial designed to determine the optimal dose and assess the efficacy and tolerability of a 6-week treatment with TG2-inhibitor ZED1227 capsules versus placebo in subjects with well-controlled CeD undergoing a gluten challenge (EU Clinical Trials Register, EudraCT Number: 2017-002241-30). The full inclusion and exclusion criteria have been published^30^. Briefly, patients had a biopsy-proven CeD diagnosis, reported a strict GFD for at least one year, were symptom-free, showed normalized duodenal histology compared to their initial diagnostic biopsy, and tested negative for TG2 antibodies at study inclusion (GFD group, Table 1). These patients were then challenged with a cookie containing 3 grams of gluten daily for 6 weeks (PGC group). Compliance of at least 80% was confirmed.

**Table 1.** Demographics and characteristics of patients.

| Variables | Drug group (n = 28) | Placebo group (n = 19) |
| --- | --- | --- |
| **Sex, no. (%)** |  |  |
| Male | 10 (35.7%) | 7 (36.8%) |
| Female | 18 (64.3%) | 12 (63.2%) |
| **Age, year** |  |  |
| Mean ± SD | 40.9 ± 15.0 | 45.3 ± 14.8 |
| Range | 22 - 65 | 19 - 62 |
| **BMI, kg/m<sup>2</sup>** |  |  |
| Mean ± SD | 25.3 ± 3.9 | 25.0 ± 4.6 |
| Range | 19.0 - 34.9 | 18.7 - 35.1 |
| **TG2 IgA, kU/L** |  |  |
| Median(Q1-Q3), at the baseline | 1 (1-2.25) | 1 (1-1.5) |
| Median(Q1-Q3), post gluten challenge | 1 (1-2) | 1 (1-7) |

**Table 2.**
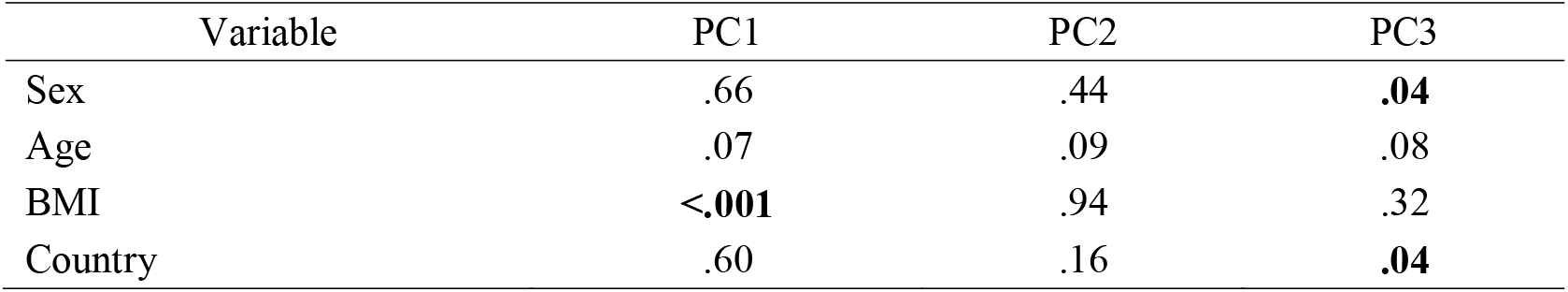
P values for the regression of Principal Components against various demographic and laboratory parameters variables. PC1, PC2, and PC3 accounted for 32.7%, 11.1%, and 7.3% of the variability, respectively, in the current data set. Significant correlations are shown in bold.

Blood samples were collected from each participant at two defined time points: at study inclusion (baseline, referred to as Gluten free diet, GFD) and at the final visit post gluten challenges (PGC) (Fig. S1). Plasma-EDTA was separated from the cell pellet and stored at −80°C.

The study analyzed samples from two groups, both before (GFD) and after the gluten challenge (PGC): the placebo arm (p) and the 100-mg ZED1227 drug arm (d) groups. The latter, being the highest dose group, showed the most significant improvement compared to the placebo and was thus selected for the current study. A total of 47 patients (drug group, n = 28; placebo group, n = 19, with a total of 94 GFD baseline and PGC samples) were included from the original 68 patients who had adequate biopsy samples at both time points^30^ and who provided separate written informed consent for these exploratory (optional) studies.

### Lipidomics

A total of 94 plasma samples were randomized and subjected to lipid extraction using a modified version of the Folch procedure.^64^ Promptly after extraction, 10 µL of 0.9% NaCl and 120 µL of CHCl_3_:MeOH (2:1, v/v) containing 2.5 µg/mL internal standard solution (for quality control and normalization purposes) were added to 10 µL of each plasma sample. A detailed protocol of plasma lipid profiling and data preprocessing is given in Supplementary Methods.

### DNA methylation

Genomics DNA was extracted from whole blood samples with Chemagic 360 robot with CMG-1091 kit (Revvity, Waltham, MA, USA) in combination with chemagic™ DNA Blood 400 kit H96 (Cat. No. CMG-1091), following the manufacture’s protocol (VD190913.che). DNA methylation profiling was performed using the Illumina Infinium MethylationEPIC version 2 BeadChip at Helmholtz Zentrum, Munich, Germany. Samples were applied to the arrays in a randomized order. Aliquots of 1 µg of DNA were subjected to bisulfite conversion, and 4 µl of bisulfite-converted DNA underwent whole-genome application, enzymatic fragmentation, and hybridization onto the MethylationEPIC BeadChip (EPICv2). The arrays were scanned using the iScan system (Illumina, San Diego, CA, USA).

### Proteomics

Plasma proteins were denatured with urea, reduced with dithiothreitol, alkylated with iodoacetamide, and then digested with trypsin in a 96-well plate format. The digests were acidified, then desalted and concentrated using C18 solid-phase extraction (SepPak C18, Waters). Samples were analysed with a Q Exactive HF Orbitrap mass spectrometer (Thermo Scientific) coupled to an Evosep One liquid chromatograph (Evosep). Further method details can be found in the Supplementary Methods.

### Statistical analysis

The statistical analysis was performed using R (Version 4.3.0 (2023-04-21)), R Foundation for Statistical Computing (Vienna, Austria).

#### Lipidomics analysis

308 lipid species were identified and included in the statistical analyses. The data underwent normalization using the autoscaling method, which involved mean centering and subsequent scaling by standard deviation. Any remaining missing values were imputed with the minimal detected expression. Significantly expressed lipids were identified using the Limma package. The model matrix included the sample group, patient BMI, age, and country as covariates. To account for patient-to-patient variability, patient ID was treated as a random effect. A p-value below 0.05, without adjustment for multiple testing, was considered statistically significant. Lipid set Enrichment analysis was conducted utilizing the fgsea package^65^ (version 1.28.0) with logFC serving as the ranking statistic for lipids, and structural classification was used for grouping information.

#### DNA methylation analysis

The DNA methylation data was pre-processed using the default meffil pipeline, with a p-value threshold of 0.05. Probes that failed quality control (QC) were removed from further analysis. Principal components were derived from control probes and estimated cell type proportions were calculated for drug and placebo groups separately using the meffil package. An epigenome-wide association study (EWAS) was performed to assess differences between baseline and post-exposure conditions, correcting the model with age, sex, the first 20 principal components and estimated cell type proportions. Analyses were performed separately for the drug and placebo groups. Comparisons of cell type proportions between the placebo and drug groups were performed at both baseline and post-exposure using Mann-Whitney U test. In addition, Spearman correlation analysis was conducted to assess CpG sites exhibiting a change in beta values greater than 2.5% in both the drug and placebo conditions across different timepoints, as well as between treatment groups at the challenge phase.

#### Proteomics analysis

Using Spectronaut software (version 19), the data were searched against the human reference proteome sequence database (20,435 entries, 26/09/2024) with the inclusion of common contaminants (381 entries^66^). Matches to the contaminants were excluded from downstream analysis. Additionally, for a protein to qualify for downstream analysis, detection of more than one unique peptide was required. Normalized protein intensity values from Spectronaut were log_2_-transformed prior to statistical analysis. Data was verified to have minimal technical variability before further analysis (Fig. S4).

Differentially expressed proteins were identified using the reproducibility-optimized test statistic (ROTS v1.34.0)^67^ with default parameters. The maximum top list size K was set to 75 % of all proteins, and the seed was set to 1. For the comparison between gluten-free diet and post-gluten challenge samples from the same placebo-treated individuals (PGCp vs GFDp), ROTS was applied in paired mode. The comparison between post-gluten challenge samples of drug-treated and placebo-treated individuals (PGCp vs PGCd) was performed using unpaired mode. P values were corrected for multiple testing using the Benjamini-Hochberg procedure^68^. To assess the overall similarity of expression changes between the two comparisons, log□ fold changes of all proteins were used to determine the Pearson correlation coefficient and its statistical significance. Statistical analyses were performed using the statistical software R version 4.4.3 (R Foundation for Statistical Computing).

## ABBREVIATIONS

CeD: Celiac disease
TG2: transglutaminase 2
GFD: gluten-free diet
GFDd: gluten-free diet drug group
GFDp: gluten-free diet placebo group
PGC: post gluten challenge
PGCd: post gluten challenge drug group
PGCp: post gluten challenge placebo group
VH:CrD: villous height to crypt depth ratio
BMI: body mass index
CAR: fatty acylcarnitines
GFR: glomerular filtration rate
HLA: human leukocyte antigen
EDTA: Ethylenediaminetetraacetic acid
TG2 IgA: anti-transglutaminase 2 immunoglobulin A antibodies
PCA: principal-component analysis
DELs: differentially expressed lipids
FA: Fatty Acyls
GP: Glycerophospholipids
ST: Sterol Lipids
SP: Sphingolipids
GL: Glycerolipids
PR: Prenol Lipids
PC O: Phosphatidylcholines
LPC: Lysophosphatidylcholines
LPE: Lysophosphatidylethanolamines
PC: Phosphatidylcholines
PE: Phosphatidylethanolamines
PE O-: Ether-linked Phosphatidylethanolamines
PI: Phosphatidylinositols
PS: Phosphatidylserines.

