## Supplemental Figures S1-S2 for "Therapeutic Restoration of Systemic Multiomic Responses by Transglutaminase 2 inhibitor in Celiac Disease: A Gluten Challenge Approach"

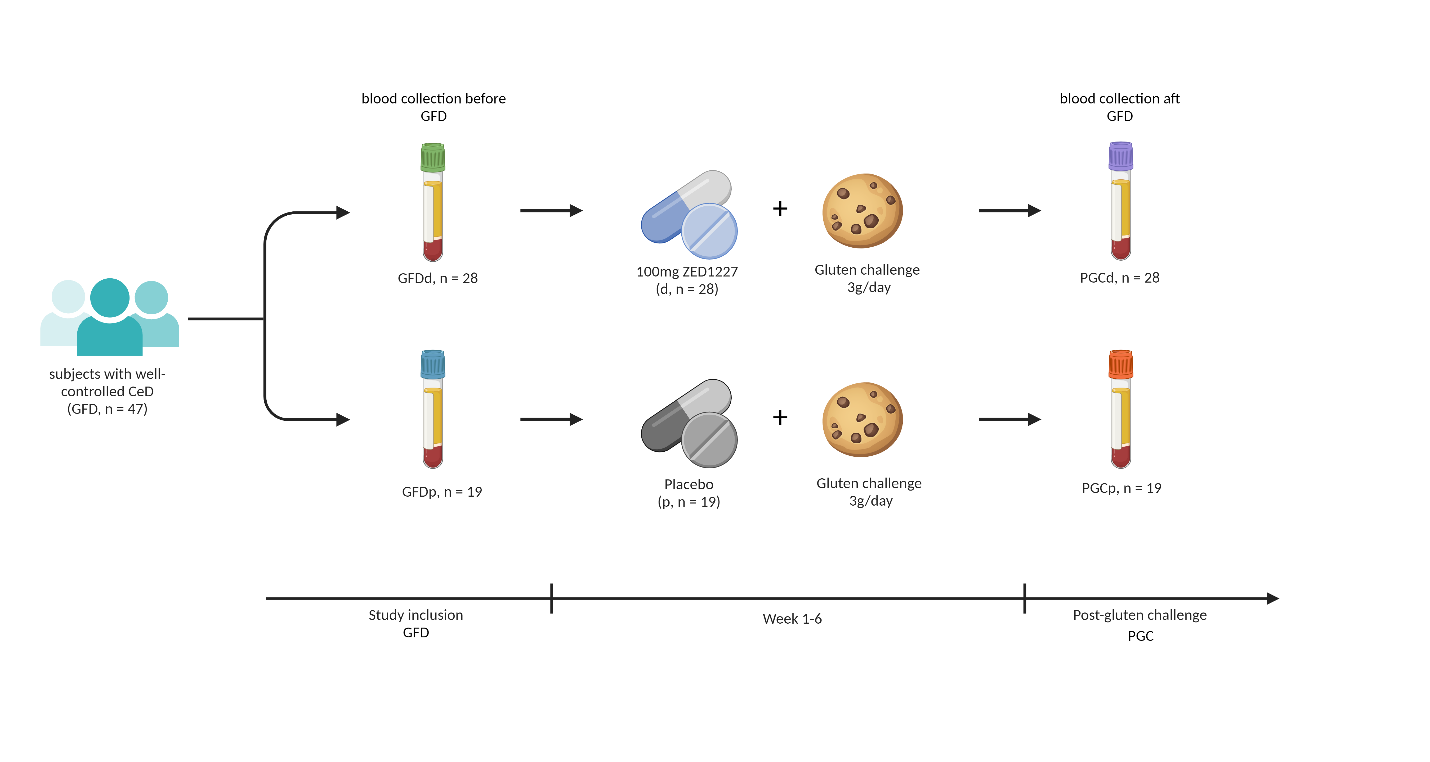

**Figure S1. Schematic presentation of the study.** Samples (n = 94; n of patients = 47), in the form of blood samples, were collected from the trial, aimed at dose-finding and assessing the efficacy and tolerability of a 6-week treatment with ZED1227 capsules vs. placebo in subjects with well-controlled celiac disease undergoing gluten challenge. Blood sampling was performed twice: on study inclusion (GFDd, n = 28; GFDp, n = 19) and at the final visit (PGCd, n = 28; PGCp, n = 19). Plasma-EDTA was separated from the cell pellet and subjected to lipidomic analysis. Created with BioRender.com.

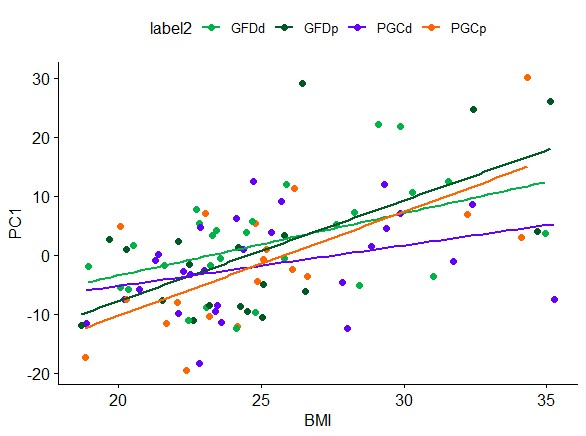

**Figure S2.** BMI trends identified by principal component analysis (PCA). Subject BMI was regressed against first principal component (PC1) scores separately for each group: GFDd (n = 28, green color), GFDp (n = 19, dark green color), PGCd (n = 28, violet color), and PGCp (n = 19, orange color). The lines of best fit for each group are colored to match the color of the symbol for that group. The violet line (for PGCd) seems to have a less steep slope compared to the other. The similarity in the slopes of the regressed lines indicates that BMI-related changes in lipidomic profiles were similar in each group and independent of drug treatment.

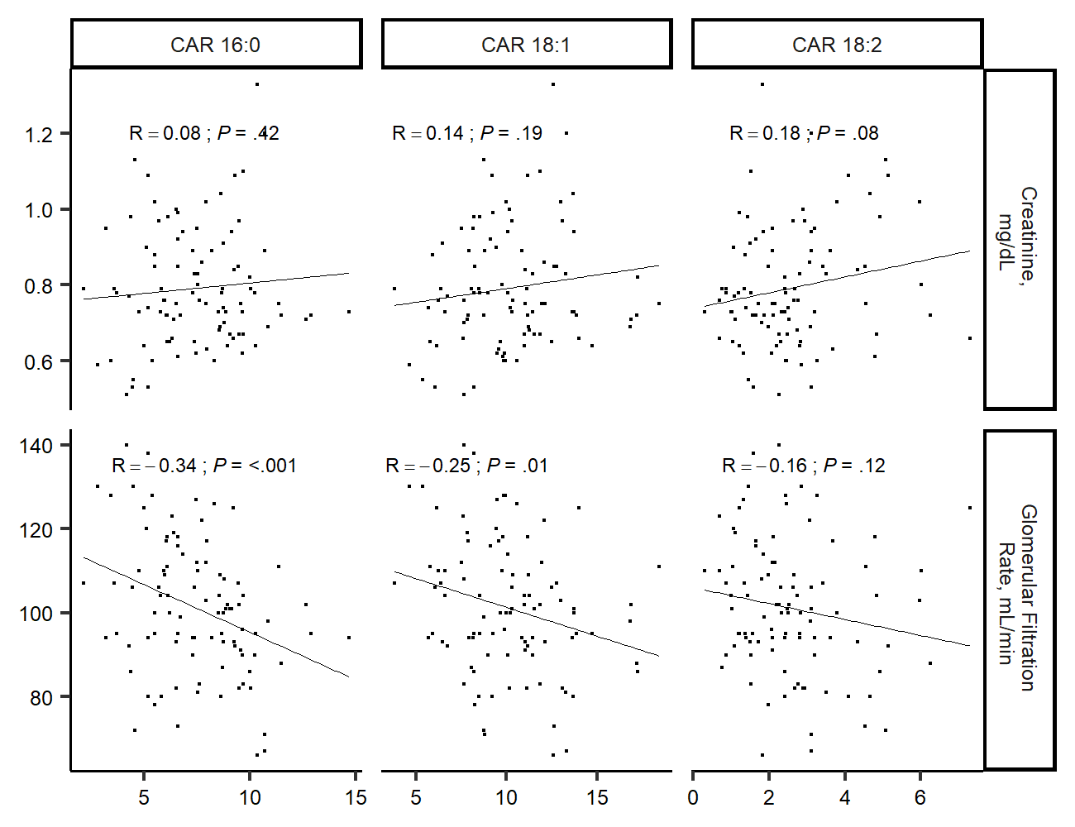

**Figure S3** Correlation of plasma Creatinine and Glomerular Filtration Rate indicators with CAR 16:0, CAR 18:1, and CAR 18:2. Pearson correlation coefficient is shown, and P-values less than 0.05 are considered to be significant. CAR - Fatty acylcarnitines; CAR 18:2 – octadecadienylcarnitine; CAR 18:1 – octadecenoylcarnitine; CAR 16:0 – Palmitoylcarnitine.

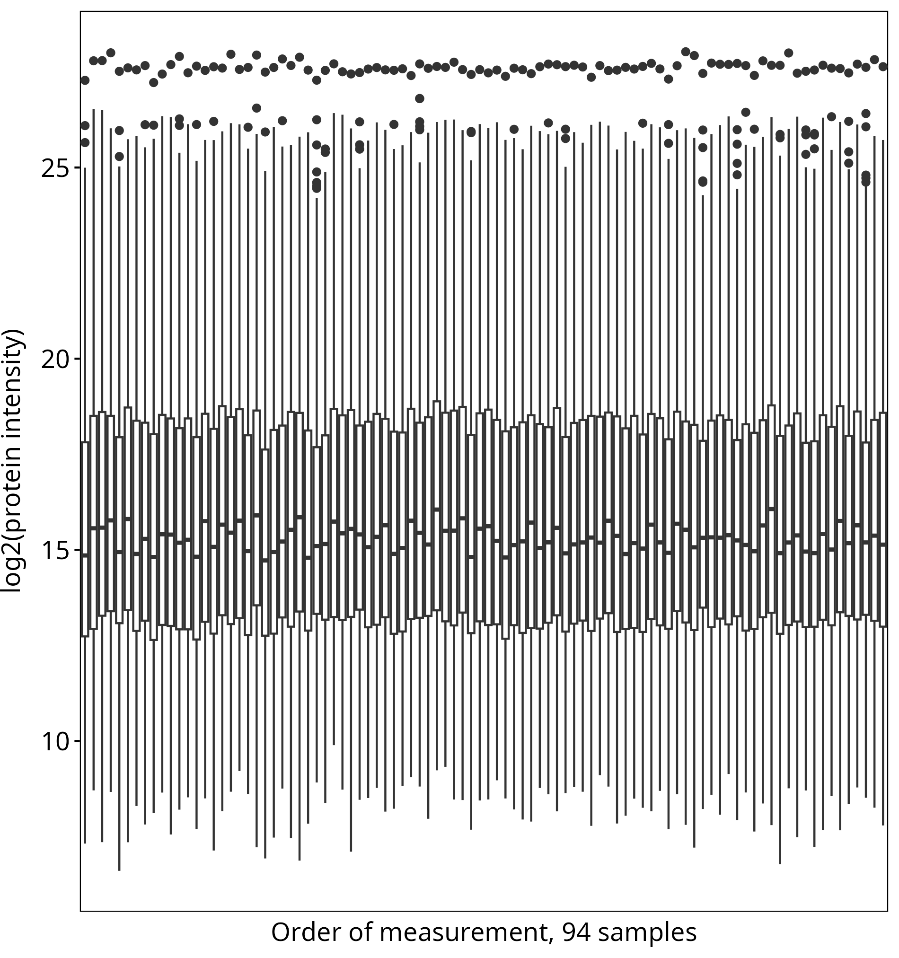

**Figure S4**. Boxplots of intensities from all proteomics samples. Stability of intensity levels suggests consistent measurement quality. All 94 samples were measured within 4 days. Whiskers of boxplots extend 1.5 times interquartile range from quantiles
