## Supplemental methods for "Therapeutic Restoration of Systemic Multiomic Responses by Transglutaminase 2 inhibitor in Celiac Disease: A Gluten Challenge Approach"

**Supplementary Methods**

**Lipidomics**

Samples were randomized and extracted using a modified version of the previously published Folch procedure (reference needed). In short, 10 µL of 0.9% NaCl and 120 µL of CHCl3: MeOH (2:1, v/v) containing the internal standards (c = 2.5 µg/mL) was added to 10 µL of each plasma sample. The standard solution contained the following compounds: 1,2-diheptadecanoyl-sn-glycero-3-phosphoethanolamine (PE(17:0/17:0)), N-heptadecanoyl-D-erythro-sphingosylphosphorylcholine (SM(d18:1/17:0)), N-heptadecanoyl-D-erythro-sphingosine (Cer(d18:1/17:0)), 1,2-diheptadecanoyl-sn-glycero-3-phosphocholine (PC(17:0/17:0)), 1-heptadecanoyl-2-hydroxy-sn-glycero-3-phosphocholine (LPC(17:0)) and 1-palmitoyl-d31-2-oleoyl-sn-glycero-3-phosphocholine (PC(16:0/d31/18:1)), (Avanti Polar Lipids, Inc., Alabaster, AL, USA), and also cholesteryl heptadecanoate (CE(17:0) and triheptadecanoylglycerol (TG(17:0/17:0/17:0)) (Larodan AB, Solna, Sweden). The samples were vortex mixed and incubated on ice for 30 min, after which they were centrifuged (9400 × g, 3 min). 60 µL from the lower layer of each sample was then transferred to a glass vial with an insert, and 60 µL of CHCl3: MeOH (2:1, v/v) was added to each sample. The samples were stored at -80 °C until analysis.

Calibration curves using 1-hexadecyl-2-(9Z-octadecenoyl)-sn-glycero-3-phosphocholine (PC(16:0e/18:1(9Z))), 1-(1Z-octadecenyl)-2-(9Z-octadecenoyl)-sn-glycero-3-phosphocholine (PC(18:0p/18:1(9Z))), 1-stearoyl-2-hydroxy-sn-glycero-3-phosphocholine (LPC(18:0)), 1-oleoyl-2-hydroxy-sn-glycero-3-phosphocholine (LPC(18:1)), 1-palmitoyl-2-oleoyl-sn-glycero-3-phosphoethanolamine (PE(16:0/18:1)), 1-(1Z-octadecenyl)-2-docosahexaenoyl-sn-glycero-3-phosphocholine (PC(18:0p/22:6)) and 1-stearoyl-2-linoleoyl-sn-glycerol (DG(18:0/18:2)), 1-(9Z-octadecenoyl)-sn-glycero-3-phosphoethanolamine (LPE(18:1)), N-(9Z-octadecenoyl)-sphinganine (Cer(d18:0/18:1(9Z))), 1-hexadecyl-2-(9Z-octadecenoyl)-sn-glycero-3-phosphoethanolamine (PE(16:0/18:1)) (Avanti Polar Lipids), 1-Palmitoyl-2-Hydroxy-sn-Glycero-3-Phosphatidylcholine (LPC(16:0)), 1,2,3 trihexadecanoalglycerol (TG(16:0/16:0/16:0)), 1,2,3-trioctadecanoylglycerol (TG(18:0/18:0/18:)) and 3β-hydroxy-5-cholestene-3-stearate (CE(18:0)), 3β-Hydroxy-5-cholestene-3-linoleate (CE(18:2)) (Larodan), were prepared to the following concentration levels: 100, 500, 1000, 1500, 2000 and 2500 ng/mL (in CHCl3:MeOH, 2:1, v/v) including 1250 ng/mL of each internal standard.

The samples were analyzed using ultra-high-performance liquid chromatography quadrupole time-of-flight mass spectrometry (UHPLC-QTOFMS). Briefly, the UHPLC system used in this work was a 1290 Infinity II system (Agilent Technologies, Santa Clara, CA, USA). The system was equipped with a multi sampler (maintained at 10°C), a quaternary solvent manager and a column thermostat (maintained at 50 °C). The injection volume was 1 µL, and the separations were performed on an ACQUITY UPLC® BEH C18 column (2.1 mm × 100 mm, particle size 1.7 µm) (Waters, Milford, MA, USA). The mass spectrometer coupled to the UHPLC was a 6545 QTOF (Agilent Technologies) interfaced with a dual jet stream electrospray (Ddual ESI) ion source. All analyses were performed in positive ion mode, and MassHunter B.06.01 (Agilent Technologies) was used for all data acquisition. Quality control was performed throughout the dataset by including blanks, pure standard samples, extracted standard samples, and control plasma samples.

Mass spectrometry data processing was performed using the open-source software package MZmine 2.53 ^53^. The following steps were applied in this processing: (i) Crop filtering with a m/z range of 350 – 1200 m/z and an RT range of 0.5 to 12 minutes, (ii) Mass detection with a noise level of 1000, (iii) Chromatogram builder with a minimum time span of 0.08 min, minimum height of 1000 and a m/z tolerance of 0.006 m/z or 10.0 ppm, (iv) Chromatogram deconvolution using the local minimum search algorithm with a 70% chromatographic threshold, 0.05 min minimum RT range, 5% minimum relative height, 1200 minimum absolute height, a minimum ration of peak top/edge of 1.05 and a peak duration range of 0.08 - 5.0, (v)~~,~~ Isotopic peak grouper with a m/z tolerance of 5.0 ppm, RT tolerance of 0.07 min, maximum charge of 2 and with the most intense isotope set as the representative isotope, (vi) Join aligner with a m/z tolerance of 0.009 or 10.0 ppm and a weight for of 2, a RT tolerance of 0.1 min and a weight of 1 and with no requirement of charge state or ID and no comparison of isotope pattern, (viii) Peak list row filter with a minimum of 10% of the samples (vii) Gap filling using the same RT and m/z range gap filler algorithm with an m/z tolerance of 0.009 m/z or 11.0 ppm, (ix) Identification of lipids using a custom database search with an m/z tolerance of 0.009 m/z or 10.0 ppm and a RT tolerance of 0.1 min, and (xii) Normalization using internal standards PE(17:0/17:0), SM(d18:1/17:0), Cer(d18:1/17:0), LPC(17:0), TG(17:0/17:0/17:0) and PC(16:0/d30/18:1)) for identified lipids and closest ISTD for the unknown lipids followed by calculation of the concentrations based on lipid-class concentration curves.

An aliquot of each sample was collected and pooled and used as a quality control sample, together with the NIST SRM1950 reference plasma sample, an in-house pooled serum sample.

**Proteomics**

Plasma protein digests were prepared in a 96-well plate format. Briefly, serum aliquots were diluted and denatured with urea, reduced with dithiothreitol, alkylated with iodoacetamide and digested with trypsin. The digests were desalted and concentrated using C18 solid-phase extraction (SepPak C18, Waters). Aliquots of the samples (800 ng) were analysed using liquid chromatography-tandem mass spectrometry with an Evosep One liquid chromatograph (Evosep) coupled to a Q Exactive HF Orbitrap (Thermo Scientific). A C_18_ reversed-phase column (15 cm x 150 µm, 1.9 µm EV-1106, Evosep Endurance) was used for the 30 sample/day method. Data were acquired using a data independent acquisition (DIA) strategy with 33 variable windows from 365 to 1005 Da. The MS1 and MS2 resolution were 120 k and 30 k, with AGCs of 3 x 10^-6^ and 1 x 10^-6^, respectively.
