## Supplementary material for "Therapeutic Restoration of Systemic Multiomic Responses by Transglutaminase 2 inhibitor in Celiac Disease: A Gluten Challenge Approach": Lict of CEC-3 investigators

**#List of CEC-3 Trial Group Collaborators**

Investigators

| **Country** | **Principal Investigator** | **Institution** |
|---|---|---|
| Estonia | **Karin Kull, MD** | Department of Gastroenterology, Internal Medicine Clinic, Tartu University Hospital, Tartu |
| Finland | **Jari Koskenpato, MD** | Lääkärikeskus Aava Helsinki Kamppi, Helsinki |
| | **Mika Scheinin, MD, PhD** | Clinical Research Services Turku - CRST Oy, Turku |
| | **Marja-Leena Lähdeaho, MD, PhD*** | Faculty of Medicine and Health Technology, Tampere University and Tampere University Hospital, Tampere<br>*and Department of Pediatrics, Tampere University Hospital, Tampere |
| Germany | | |
| | **Michael Schumann, MD** | Department for Gastroenterology, Infectious diseases and Rheumatology, Campus Benjamin Franklin, Charité - University Medicine Berlin, Berlin |
| | **Yurdagül Zopf, MD** | Department of Medicine 1, Hector Center for Nutrition, Exercise, and Sports, Universitätsklinikum Erlangen, Friedrich-Alexander-University Erlangen-Nürnberg, Erlangen |
| | **Andreas Stallmach, MD** | Department of Internal Medicine IV, Jena University Hospital, Friedrich-Schiller University Jena, Jena |
| | **Ansgar W. Lohse, MD** | I. Department of Medicine, University Medical Center Hamburg-Eppendorf, Hamburg |
| | **Stefano Fusco, MD** | Division of Gastroenterology, Hepatology, Infectious Diseases, Department of Internal Medicine I, University Hospital Tübingen, Tübingen |
| Germany | **Jost Langhorst, MD** | Department for Internal and Integrative Medicine, Kliniken Essen-Mitte, Essen |
| | **Jost Langhorst, MD** | Department for Internal and Integrative Medicine, Sozialstiftung Bamberg, Chair for Integrative Medicine, University of Duisburg-Essen, Bamberg |
| | **Helga Paula Török, MD** | Department of Medicine II, University Hospital, LMU Munich, Munich |
| Ireland | **Valerie Byrnes, MD** | University College Hospital Galway, Galway |

| Country | Principal Investigator | Institution |
|---|---|---|
| Lithuania | **Juozas Kupcinskas, MD** | Gastroenterology Department and Institute for Digestive Research, Lithuanian University of Health Sciences, Kaunas |
| Norway | | |
| | **Øistein Hovde, MD, PhD** | Medical Department, Innlandet Hospital Trust, Gjøvik |
| | **Jørgen Jahnsen, MD** | Akershus University Hospital, Lørenskog |
| Switzerland | **Luc Biedermann, MD** | Department of Gastroenterology and Hepatology, University Hospital Zürich, Zürich |
| | **Jonas Zeitz, MD** | Swiss Celiac Center, Center of Gastroenterology, Clinic Hirslanden, Zürich and Department of Gastroenterology and Hepatology, University Hospital Zürich, Zürich |
